# Patterns of antivenom administration among snakebite patients in selected Zambian hospitals: a retrospective multi-facility review

**DOI:** 10.64898/2026.08.07.26358265

**Authors:** Kingford Chimfwembe, Aashna Uppal, Paul Kingpriest, Marcel van Driel, Frank Shamilimo, Namasiku Siyumbwa, Arnold Hamapa, Mweetwa Mudenda, Frank Tianyi, Andreas Deckert, Hans Joerg-Lang, Ulrich Kuch, James Mwansa, Lillian Mambwe Mutesu, Lydia Hangulu

## Abstract

**Background:** Snakebite envenoming remains a major but neglected public health problem in sub-Saharan Africa, where access to antivenom is often limited and unevenly distributed. In Zambia, evidence on facility-level determinants of antivenom use is scarce. This study examined factors associated with antivenom administration among snakebite patients in selected health facilities.

**Methods:** A retrospective cross-sectional study was conducted among 128 snakebite patients presenting to five health facilities in Zambia. Associations between antivenom administration and health facility, admission year, age group, and gender were assessed using Fisher’s exact test, with statistical significance set at p < 0.05.

**Results:** Overall, 16/128 patients (12.5%) received antivenom. Antivenom administration varied significantly by health facility (p < 0.0001). All patients at Kabwe General Hospital received antivenom (9/9, 100%), compared to 50% (2/4) at Commando Urban Health Centre and 22.7% (5/22) at Chipata Central Hospital. No patients received antivenom at Mpongwe Mission Hospital (0/70) or St. Dominic’s Mission Hospital (0/23), where antivenom was not available. Admission year (2023 vs 2024; p = 0.78), age group (p = 0.53), and gender (p = 0.80) were not significantly associated with antivenom use.

**Conclusion:** Antivenom administration in this setting is determined primarily by health facility-level availability rather than patient demographic or temporal factors. The very low overall treatment rate highlights critical inequities in access to life-saving therapy. Strengthening antivenom procurement, equitable distribution, and referral systems is urgently needed to reduce preventable morbidity and mortality from snakebite envenoming.

## Introduction

Snakebite envenoming remains a substantial global public health problem. Early landmark estimates suggested that at least 421,000 envenomings and 20,000 deaths occurred annually worldwide, with possible upper estimates reaching 1.84 million envenomings and 94,000 deaths (1). More recent estimates however, indicate that approximately 5.4 million snakebites occur annually, leading to 1.8-2.7 million envenomings and between 81,000 and 138,000 deaths each year (2,3). In addition, nearly three times as many survivors experience amputations and other permanent disabilities(4,5).

Globally, approximately 5.8 billion people are considered at risk of snakebite envenoming, with an estimated 7,400 bites occurring daily and resulting in 220 - 380 deaths per day (1,6). Further, the pooled global incidence of snakebite envenoming is estimated at 69.4 per 100,000 population annually, while pooled mortality is approximately 0.33 per 100,000 population per year (2). In 2019 alone, the Global Burden of Disease (GBD) estimated 63,400 snakebite-related deaths globally, with an age-standardized mortality rate of 0.8 deaths per 100,000 population (7). Although this represented a 36% decline in mortality since 1990, current progress remains insufficient to achieve the WHO 2030 targets(8).

In terms of global distribution, the highest burden of snakebite exists in South Asia, Southeast Asia, and sub-Saharan Africa, estimating at least 421,000 envenomings and 20,000 deaths annually worldwide, with upper bounds reaching 1,841,000 envenomings and 94,000 deaths (1). In sub-Saharan Africa specifically, where SBE data is still a challenge, up to one million people are reported as being bitten each year, with estimates of 7,00020,000 deaths annually (9). The annual burden of SBE has been estimated at 1.03 million DALYs mainly from snakebite-envenoming-related deaths, amputations, and post-traumatic stress disorder, which is similar to or higher than the burden of many other NTDs (9). More recent estimates however, suggest snakebites in sub-Saharan Africa account for 20,000 to 32,000 annual deaths, though these numbers are likely underestimated due to reliance on hospital data and incomplete central databases (10). Approximately 435,000 to 580,000 snakebite victims in Africa require medical attention each year. Incidence estimates vary widely across the continent, ranging from about 5 to over 400 per 100,000 population depending on region and rurality. Mortality usually ranges from 2% to 3%, though this figure may not present the true picture due to significant underreporting and treatment gaps(11).

Snakebite envenoming remains a significant public health challenge in Zambia, particularly in rural areas where agricultural activities predominate and access to healthcare services is often limited (12). Epidemiological data on snakebite in Zambia remain fragmented and incomplete, reflecting broader challenges in snakebite surveillance across sub-Saharan Africa (13). According to the World Health Organization, one consequence of the historically inadequate prioritization and control of snakebite envenoming has been the persistent lack of high-quality epidemiological data, resulting in surveillance systems that often lack both completeness and geographical resolution (WHO, 2019). The accuracy of available estimates is further compromised by low healthcare-seeking behaviour among snakebite victims, many of whom rely on traditional healers and other alternative forms of treatment rather than presenting to health facilities (13). Consequently, substantial under-reporting occurs, with some countries in the region estimated to miss more than 70% of snakebite cases, particularly in rural and underserved areas with limited infrastructure (13).

Until recently, reliable and up-to-date data on the burden of snakebite in Zambia were lacking. Consequently, estimates of the national burden were derived through extrapolation from community-based studies conducted in comparable settings. In particular, findings from a rural community survey in Mozambique were used to estimate the incidence of snakebite in Zambia based on similarities in rural population characteristics and ecological conditions. These estimates suggested that approximately 36,519 Zambians experience snakebites annually, resulting in an estimated 4,719 deaths each year. Furthermore, only 16.8% of snakebite victims were estimated to seek care at health facilities, indicating that the majority of cases occur outside the formal healthcare system. As a result, deaths recorded in health facilities represent only a small proportion of the overall mortality burden, with an estimated 343 deaths occurring in health centres, accounting for just 7.3% of all snakebite-related deaths.(10).

More recently, the availability of routine health information through Zambia’s District Health Management Information System (DHMIS) has provided additional insights into the epidemiology of snakebite and the burden presenting to the formal healthcare system. Although facility-based data do not capture all snakebite cases occurring in the community, they offer valuable information on temporal and geographical patterns of healthcare utilization for snakebite. DHMIS records indicate that 98,250 snakebite cases were reported nationally between 2020 and 2025, highlighting the substantial burden of snakebite among patients seeking care. Provincial data show that Central Province recorded the highest number of reported cases (14,361), followed by Copperbelt Province (13,638). Similarly high case counts were observed in Eastern Province (12,410) and Northern Province (12,203), suggesting that these regions represent important hotspots for snakebite.

Antivenom is the only specific treatment for snakebite envenoming. Monovalent antivenoms are typically administered to victims of venomous snakebites when the species responsible is known, whereas polyvalent antivenoms or combinations of multiple monovalent antivenoms are often required when envenomation is caused by an unidentified venomous snake(14). Conventional antivenoms based on animal-derived polyclonal antibodies remain the cornerstone of treatment, alongside supportive medical and surgical care (15).

Globally, 68.3% of all snakebite envenoming (SBE) cases receive hospital treatment, and 64.7% are treated with antivenom, while approximately 57% of snakebites result in envenoming (16). These figures underscore the need for an integrated global approach to snakebite envenoming, recognizing that an effective healthcare response requires coordinated efforts across multiple disciplines, including toxicology, epidemiology, and clinical medicine (17–20). However, access to antivenom remains a critical barrier to effective snakebite management, particularly in Africa. As the region with the highest incidence and burden of snakebite, achieving the World Health Organization’s goal of halving the global burden of snakebite by 2030 will require substantial improvements in antivenom access, especially in the most affected areas (21).

Current evidence from Africa reveals considerable variation in access to antivenom. For example, in Kenya, antivenom is available in approximately 45% of public health facilities and fewer than 20% of private facilities (12). The situation is even more concerning in Uganda, where only 4% of health facilities across the public, mission, and private sectors stock antivenom. This limited availability persists despite increasing morbidity and mortality associated with SBE, while the high cost of antivenom renders it unaffordable for many patients (12). In Zambia, significant gaps in antivenom availability have also been reported across public, mission, and private health facilities (22).

The objective of this study was to determine the prevalence of antivenom administration among snakebite patients treated at selected health facilities in Zambia during 2023 and 2024 and to identify patient- and facility-level factors associated with antivenom use. We hypothesised that antivenom administration would vary according to health facility characteristics and availability of antivenom stocks.

## Methods

### Study setting

The study was conducted in three provinces of Zambia, Central, Copperbelt, and Eastern and included five health facilities located within these provinces. Data were collected between May and July 2025. The provinces and health facilities were purposively selected based on the high burden of snakebite cases reported through the District Health Management Information System (DHMIS) during 2023 and 2024.

In Central Province, data were collected from Kabwe General Hospital in Kabwe District. In Copperbelt Province, data collection was undertaken in two districts: Ndola District, where Commando Urban Hospital and St. Dominic’s Mission Hospital were included, and Mpongwe District, where data were collected from Mpongwe Mission Hospital. In Eastern Province, data were collected from Chipata Central Hospital and Gondar Urban Health Centre in Chipata District.

### Study design

The study employed a retrospective review of hospital records for patients presenting with snakebite. In each selected health facility, the hospital records team, in collaboration with the principal investigator, reviewed patient records to identify eligible snakebite cases. As a retrospective review, all eligible snakebite records meeting the inclusion criteria within the study period were included. No formal sample size calculation was performed. Records meeting the predefined inclusion criteria were selected for data extraction. Eligible records included snakebite cases documented between 1 January 2023 and 31 December 2024 and containing information for at least 50% of the study variables of interest.

### Data collection

Data were collected using a structured data extraction tool developed to capture relevant information from patient records of snakebite cases. The tool collected data on the health facility, year of presentation, patient demographic characteristics (age and sex), and antivenom administration status.

To minimise information bias, data extraction was performed using a standardized data extraction tool and records were reviewed jointly by trained records personnel and the principal investigator. Data quality checks were conducted by two independent analysts, and unclear information was verified against original records where available.

Several measures were implemented to minimise potential sources of bias. A standardized data extraction tool was used across all participating facilities to ensure consistency in data collection. Data extraction was conducted in collaboration with hospital records personnel familiar with the record systems. To minimise information bias arising from incomplete or inaccurate documentation, extracted data were verified against original patient records where available. Furthermore, two independent data analysts conducted data quality checks to identify and correct inconsistencies, thereby improving the accuracy and reliability of the dataset.

### Data management and analysis

Data entry and cleaning were performed using Microsoft Excel. To ensure data quality, two independent data analysts conducted consistency and accuracy checks on all variables extracted from hospital records. Missing, incomplete, or unclear information was cross-checked against original patient records where available. Variables were updated when verification was possible and coded as missing when verification could not be completed.

The cleaned dataset was subsequently imported into the Statistical Package for the Social Sciences (SPSS) for analysis. Records with more than 50% missing information on study variables were excluded from the analysis in accordance with the study inclusion criteria. For records included in the study, missing or unclear information was verified against original patient records where available. Where verification was not possible, the variable was coded as missing. Given the low proportion of missing data among included records, analyses were conducted using complete-case analysis, with no imputation of missing values.

Age was extracted as a continuous variable from patient records and subsequently categorised into age groups (0–5 years, 6–12 years, 13–17 years, 18–39 years, 40–59 years, 60–79 years, and 80 years and above) for descriptive analysis. For inferential analysis, age categories were further grouped into children (<13 years), adolescents (13–17 years), and adults (≥18 years) to facilitate statistical comparison and ensure adequate cell counts for Fisher’s exact test. Descriptive statistics were used to summarize the study variables. As all variables were categorical, results were presented as frequencies and percentages. Age was categorised into clinically relevant groups (children, adolescents and adults) to facilitate interpretation and because of the limited sample size available for inferential analyses. Subgroup analyses were conducted to assess differences in antivenom administration across patient and facility characteristics. Fisher’s exact test was used to examine associations between categorical variables because the assumptions required for the chi-square test were not met. Statistical significance was assessed at a p-value of <0.05.

### Ethical considerations

Ethical approval for the study was obtained from the Lusaka Apex Medical University Biomedical Research Ethics Committee (LAMUBREC) (Ref: LAMUBREC No. 0534/13/03/2025; approved on 13 March 2025) and the National Health Research Authority (NHRA), Zambia (Ref: NHRA-2144/17/04/2025; approved on 22 April 2025). Additional administrative permission was obtained from the relevant provincial and district health offices, as well as from the participating health facilities prior to data collection.

## Results

### Study participant selection

A total of 164 snakebite records were identified from the participating health facilities for the period 1 January 2023 to 31 December 2024. Of these, 36 records were excluded because they contained less than 50% of the required study variables or had insufficient information for analysis. The remaining 128 records met the eligibility criteria and were included in the final analysis.

### Characteristics of the snakebite Victims

A total of 128 snakebite patients were included in the analysis. The sample characteristics are summarised in Table 1. The majority of patients were from Mpongwe District (54.7%, n = 70) and were admitted to Mpongwe Mission Hospital (54.7%, n = 70). Most patients presented in 2023 (60.9%, n = 78) compared with 2024 (39.1%, n = 50). Adults aged ≥18 years comprised the largest age group (61.7%, n = 79), followed by children under 12 years (27.3%, n = 35) and adolescents aged 13–17 years (10.9%, n = 14). There was a slight female predominance (53.1%, n = 68) compared with males (46.9%, n = 60).

**Table 1:** Sample Characteristics.

| Variable | Category | n | % |
| --- | --- | --- | --- |
| **District** |  |  |  |
|  | Kabwe | 9 | 7.0 |
|  | Mpongwe | 70 | 54.7 |
|  | Ndola | 27 | 21.1 |
|  | Chipata | 22 | 17.2 |
| **Health Facility** |  |  |  |
|  | Kabwe General Hospital | 9 | 7.0 |
|  | Mpongwe Mission Hospital | 70 | 54.7 |
|  | St. Dominic Mission Hospital | 23 | 18.0 |
|  | Commando Urban Centre | 4 | 3.1 |
|  | Chipata Central Hospital | 22 | 17.2 |
| **Admission Year** |  |  |  |
|  | 2023 | 78 | 60.9 |
|  | 2024 | 50 | 39.1 |
| **Age Group** |  |  |  |
|  | Children <12 years | 35 | 27.3 |
|  | Adolescents (13–17 years) | 14 | 10.9 |
| | Adults ($\geq 18$ years) | 79 | 61.7 |
| **Gender** |  |  |  |
|  | Female | 68 | 53.1 |
|  | Male | 60 | 46.9 |

### Administration of antivenom

The data shows a very low rate of antivenom administration in the selected health facilities. Out of a total of 128 cases, only 16 (approximately 12.5%) received antivenom, while the vast majority, 112 cases (about 87.5%), did not.

**Figure 1:**
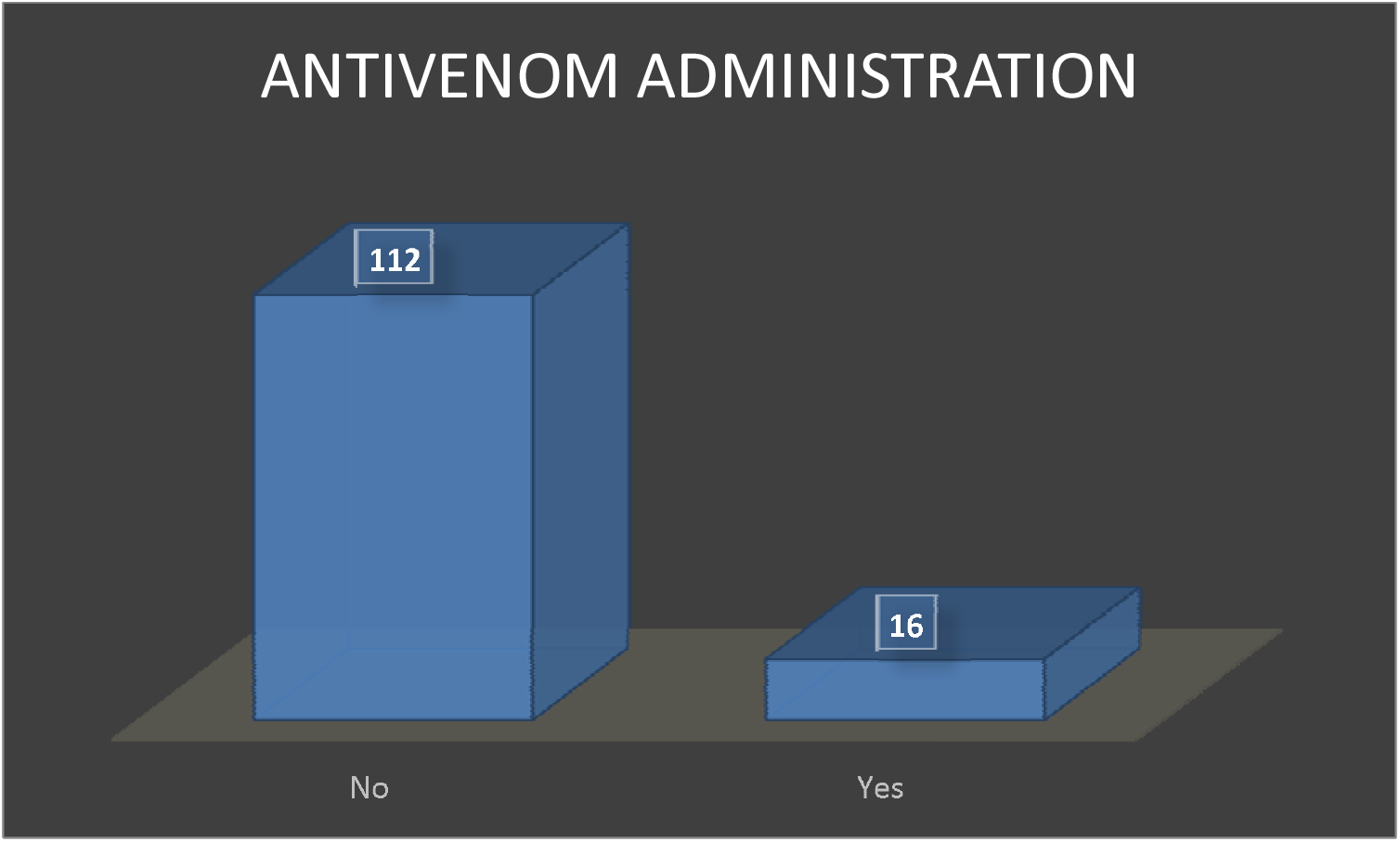
Administration of antivenom.

### Availability of Antivenom at the Time of the Facility Visits

The availability of antivenom varied considerably across the study sites. During the time of data collection, antivenom was stocked only at Chipata Central Hospital, Kabwe General Hospital, and Commando Urban Health Centre in Ndola. In contrast, Mpongwe Mission Hospital and St. Dominic’s Mission Hospital did not have any antivenom in stock.

### Factors Associated with Antivenom Administration

#### Health facility and district

Antivenom administration varied significantly by health facility (χ^2^ = 83.53, df = 4, P < 0.001) and by district (χ^2^ = 75.74, df = 3, P < 0.001). All nine patients (100%) admitted to Kabwe General Hospital received antivenom, compared with none of the 70 patients (0%) at Mpongwe Mission Hospital and none of the 23 patients (0%) at St. Dominic Mission Hospital. At Commando Urban Centre, 2 of 4 patients (50.0%) received antivenom, while at Chipata Central Hospital, 5 of 22 patients (22.7%) received antivenom. Similarly, all nine patients (100%) from Kabwe District received antivenom, compared with none from Mpongwe District (0%, n = 70), 2 of 27 from Ndola District (7.4%), and 5 of 22 from Chipata District (22.7%). Due to small expected cell counts (<5 in 50% of cells for health facility and 37.5% for district), Fisher’s exact test was used to confirm these associations.

#### Admission year

There was no significant association between admission year and antivenom administration (χ^2^ = 0.17, df = 1, P = 0.68; Fisher’s exact test, P = 0.79). Among patients admitted in 2023, 9 of 78 (11.5%) received antivenom, compared with 7 of 50 (14.0%) in 2024.

#### Age group

Age group was not significantly associated with antivenom receipt (χ^2^ = 2.80, df = 2, P = 0.25). Antivenom was administered to 7 of 35 children (20.0%), 2 of 14 adolescents (14.3%), and 7 of 79 adults (8.9%).

#### Gender

There was no significant association between gender and antivenom administration (χ^2^ = 0.07, df = 1, P = 0.79; Fisher’s exact test, P = 1.00). Antivenom was received by 9 of 68 females (13.2%) and 7 of 60 males (11.7%).

**Table 2:**
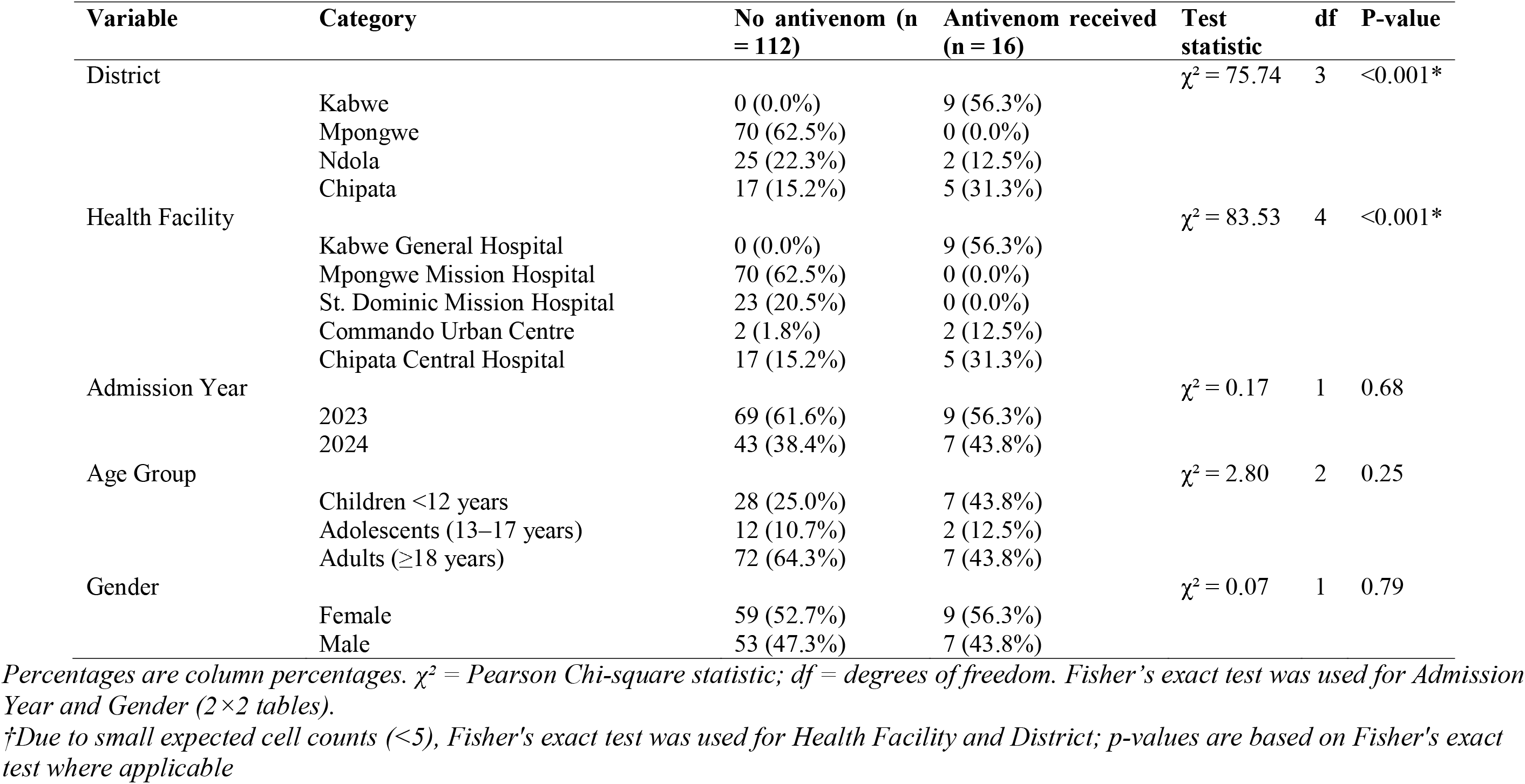
Bivariate analysis of factors associated with antivenom administration among snakebite patients (N = 128)

## Discussion

This study examined factors associated with antivenom administration among snakebite patients presenting to selected health facilities in Zambia. The key finding is that only 12.5% of patients received antivenom, with health facility being the only statistically significant determinant of administration, while age, gender, and admission year were not significant predictors. These findings suggest that access to antivenom is driven predominantly by health system factors, particularly facility-level availability and capacity, rather than patient demographic characteristics.

The overall rate of antivenom administration observed in this study is markedly low. Despite being the only specific and potentially life-saving treatment for venomous snakebite, only a small proportion of patients received antivenom. This reflects persistent systemic challenges in ensuring timely access to appropriate care. Limited access to safe and effective antivenoms is a major barrier to reducing snakebite-related morbidity and mortality globally (2,23,24). Similar challenges have been documented across sub-Saharan Africa, where supply chain inefficiencies, weak procurement systems, inadequate forecasting, and constrained national health budgets continue to limit availability ((6,17,25,26).

A particularly important finding is the strong influence of health facility on treatment outcomes. All patients presenting to Kabwe General Hospital received antivenom, whereas none of those presenting to Mpongwe Mission Hospital or St. Dominic’s Mission Hospital received it. This pattern closely reflects antivenom availability during the study period, where only selected facilities had stock. Consequently, access to definitive treatment was largely determined by the facility attended rather than clinical need, highlighting a critical structural inequity in service delivery. Similar patterns have been reported in Nigeria, where facility readiness, including antivenom availability and provider expertise, significantly determines treatment provision ((26,27).

The observed disparities also underscore broader weaknesses in health system preparedness. Facilities such as Mpongwe Mission Hospital, which accounted for a substantial proportion of snakebite cases, lacked antivenom entirely, indicating a mismatch between disease burden and resource allocation. This is particularly concerning in rural, agriculture-dependent settings where snakebite risk is high. The global strategy for snakebite envenoming emphasizes the need for equitable distribution of antivenom based on epidemiological burden to reduce preventable deaths (28,29) .The findings of this study suggest that current distribution mechanisms may not adequately reflect local burden or service demand.

The lack of a statistically significant association between admission year and antivenom administration suggests that access to treatment remained largely unchanged between 2023 and 2024. Although there was a slight increase in utilization over time, this was not meaningful, indicating limited progress in strengthening procurement systems or improving clinical service delivery. This stagnation is consistent with evidence from other African settings where snakebite has historically received limited policy attention and funding relative to other neglected tropical diseases (9,30).

Similarly, neither age nor gender was significantly associated with antivenom administration. While children showed a higher proportion of antivenom receipt, this difference was not statistically significant. This may reflect clinical caution among healthcare providers, as children are more vulnerable to severe envenoming due to higher venom-to-body weight ratios (31–33). However, the small number of antivenom cases limits the ability to draw strong conclusions. The absence of gender differences suggests that once patients reach health facilities, treatment decisions are primarily guided by clinical presentation rather than sex-based disparities, consistent with findings from other African studies (34,35).

Although the study was conducted in only five health facilities across three provinces, the selected facilities were located in areas reporting a high burden of snakebite and represented both hospital and mission health facility settings. Therefore, the findings are likely to reflect challenges in antivenom availability and access experienced in other high-burden areas of Zambia. However, caution should be exercised when generalising the findings to all health facilities nationally, particularly those in provinces not included in the study. Further studies involving a larger and nationally representative sample of health facilities are warranted.

### Implications

The findings of this study have important implications for health policy and system strengthening. First, antivenom procurement and distribution systems should be urgently restructured to ensure alignment between stock availability and local snakebite burden. Facilities with high caseloads but no antivenom should be prioritised. Second, referral systems must be strengthened to ensure rapid transfer of patients from non-stocked facilities to treatment centres with antivenom availability. Third, continuous professional development should be enhanced to improve healthcare workers’ competence and confidence in snakebite management. Finally, routine health information systems should incorporate detailed snakebite indicators to support monitoring of treatment access, utilisation patterns, and outcomes.

Although the study included facilities from three provinces with a high burden of snakebite, the purposive selection of facilities may limit generalisability to all health facilities in Zambia. Nevertheless, the findings provide important insights into antivenom access in settings with substantial snakebite burden and may be relevant to similar resource-constrained contexts.

### Limitations

This study is limited by its retrospective design and absence of clinical severity data, which likely influences antivenom decisions. The small number of treated cases also limits statistical power for subgroup analyses. Nonetheless, the consistency of facility-level effects provides strong signal of structural inequity.

## Conclusion

Overall, the findings demonstrate that antivenom administration in the study setting is determined primarily by facility-level availability rather than patient demographic characteristics. The low treatment rate and marked inter-facility disparities highlight significant gaps in equitable access to lifesaving care. Addressing these gaps will require coordinated efforts to strengthen procurement systems, improve distribution strategies, enhance health worker capacity, and reinforce referral networks to ensure timely and equitable access to antivenom for all snakebite patients.

## Data Availability

All data produced in the present work are contained in the manuscript

## Author Contributions

KC conceived the study, developed the study design, led data collection, and led manuscript drafting. AU contributed to data cleaning, preliminary analysis and interpretation of findings. AD contributed to and critical revision of the methodology of the manuscript .PK, FS, NS, AH, MM, JM, LMM, HL and UK contributed to editing the manuscript. LH supervised the entire research process. All authors read and approved the final manuscript and agree to be accountable for all aspects of the work.

## Acknowledgments

The authors would like to acknowledge the management and staff of the participating health facilities for their support during data collection. We also extend our gratitude to all individuals who contributed to the successful completion of this study.

